# Creatine-weighted imaging in patients with Parkinson’s disease

**DOI:** 10.1101/2025.06.15.25329644

**Authors:** Kexin Wang, Nirbhay Narayan Yadav, Zijiang Yang, Ted M. Dawson, Peter van Zijl, Kelly A. Mills, Jiadi Xu, Jannik Prasuhn

## Abstract

**Background:** Parkinson’s disease (PD) is a progressive neurodegenerative disorder involving impaired bioenergetics and mitochondrial dysfunction. Creatine (Cr) supplementation has been suggested as a pathophysiology-targeted therapy, yet human studies have yielded heterogeneous results. This study employs guanidino chemical exchange saturation transfer (GuanCEST) magnetic resonance imaging (MRI), a novel Cr-weighted imaging technique, to evaluate Cr level changes in patients with PD (PwPD) compared to healthy controls (HCs).

**Methods:** 25 PwPD and 24 age- and sex-matched HCs underwent standardized clinical assessments and GuanCEST MRI. Region-of-interest (ROI) and voxel-wise analyses were conducted to assess group differences. Kendall’s correlation and ANCOVA were used to explore associations between GuanCEST signals, disease presence, and clinical severity.

**Results:** GuanCEST signals in the caudate nucleus were significantly lower in PwPD (1.67 ± 0.26%) than in HCs (1.82 ± 0.16%; p = 0.023). Signal reduction correlated with increasing PD severity, particularly in thalamic subregions. In the internal medullary lamina, GuanCEST values negatively correlated with MDS-UPDRS-III scores (r = −0.44, p = 0.03) and a trend was also seen in the lateral thalamic nuclei (r = −0.39, p = 0.06). ANCOVA indicated GuanCEST values in the internal medullary lamina decreased by ∼0.01% per point increase in MDS-UPDRS-III (p = 0.007), adjusted for age and sex.

**Conclusion:** GuanCEST MRI shows promise as a non-invasive tool for detecting Cr alterations in PD. This technique may enhance our understanding of Cr metabolism in PD and support the development of targeted therapeutic strategies.

## INTRODUCTION

Parkinson’s disease (PD) is a progressive neurodegenerative movement disorder with a complex etiology [1]. While the mechanisms underlying PD are not fully understood, a hallmark feature of the disease is the loss of dopaminergic neurons, particularly in the substantia nigra pars compacta (SNc) [2]. The selective vulnerability of these neurons has been extensively studied, with prevailing theories suggesting their high energy demands may make them more susceptible to energy supply disruptions [3]. SNc dopaminergic neurons are known to form larger axonal arbors and more synapses than other dopaminergic neurons, leading to a pronounced redistribution of mitochondria to their axonal terminals. This, in turn, elevates their energy requirements and increases susceptibility to factors that impair energy supply [4]. Furthermore, a growing body of evidence implicates impaired bioenergetics, primarily due to mitochondrial dysfunction, as a critical feature of PD pathophysiology [5,6].

Creatine (Cr) plays an essential role as a short-term energy buffer, working alongside phosphocreatine (PCr) in the phosphocreatine shuttle system to regulate cellular energy metabolism, especially in the central nervous system where mitochondria are key players in energy production [7]. During periods of heightened metabolic demand, PCr donates a phosphate group to adenosine diphosphate (ADP), rapidly replenishing adenosine phosphate (ATP) stores and converting to Cr [8]. This PCr-ATP shuttle sustains neuronal function independently of oxygen levels, which is crucial during metabolic stress [9]. Furthermore, Cr also appears to have additional roles beyond energy buffering, including antioxidant, immunomodulatory, and neuromodulatory functions (e.g., modulating GABAa receptors and glutamatergic transmission via NMDA receptors) [10,11]. Many of these effects may be linked to its bioenergetic role within mitochondria [12–14], making Cr a promising pathophysiology-orientated biomarker for mitochondrial dysfunction in PD. Currently, no medical imaging modality can directly measure Cr in the brain non-invasively. Alternative methods include 1H magnetic resonance spectroscopy (MRS), which quantifies total Cr (tCr) as the sum of Cr and PCr, and 31P MRS, which detects phosphorus-containing metabolites such as PCr and ATP, but both MRS methods suffer from poor spatial resolution. A 1H MRS study of 20 PD patients reported a significant decrease in tCr in the putamen [15]. Additionally, a meta-analysis of 31P MRS in PD (n = 183) revealed a reduction in ATP/Pi, with more pronounced decreases in regions such as the midbrain, suggesting impaired energy homeostasis as a key factor [16]. However, high-resolution, non-invasive imaging methods for *in-vivo* Cr assessment remain technically challenging and have not yet been a standard-of-care approach in the diagnostic workup of patients with PD (PwPD).

Chemical exchange saturation transfer (CEST) is a non-invasive MRI technique that enables the detection of low-concentration solutes *in vivo* without the use of contrast agents [17]. It leverages the exchange of protons between solutes and water, measuring changes in the saturated water signal as an indirect indicator of metabolite presence. A commonly used analysis method, magnetization transfer ratio asymmetry (MTR_asym_) [18], has been employed to study solutes such as mobile proteins and peptides, as in amide proton transfer-weighted (APTw) imaging. APTw MRI has been applied to various conditions, including cancer [19], Parkinson’s disease (PD), and Alzheimer’s disease (AD) [20]. However, recent studies have shown that the APTw signal includes contributions beyond amide protons, such as the amide Nuclear Overhauser Effect (amideNOE) [21,22]. In contrast, the amideCEST peak—originating from unstructured proteins [23,24]—offers greater specificity and pH sensitivity than the broader APTw signal. Cr, which contains guanidinium protons, was historically considered undetectable in CEST imaging at clinical field strengths (e.g., 3T) due to its rapid exchange rate in phantom studies [25]. More recent in vivo studies, however, have demonstrated a slower exchange rate, allowing Cr detection at 3T [26,27]. Arginine, found in mobile proteins, also contributes guanidinium protons [28]. As a result, both Cr and arginine contribute to the guanidino CEST (GuanCEST) signal [29,30].

In this study, we applied GuanCEST imaging to PwPD and age- and sex-matched HCs to assess Cr-weighted signal changes. We also incorporated amideCEST to help differentiate CrCEST from ArgCEST contributions. To our knowledge, this is the first 3T application of Cr-weighted mapping in PwPD, with a comprehensive analysis of its association with clinical disease severity.

## METHODS

### Recruitment and clinical characteristics

The present study has been performed after receiving prior approval from the institutional review board of Johns Hopkins University, Baltimore, MD, USA, (#00371335) following the revised version of the Declaration of Helsinki. All participants gave their informed written consent before any study-related activities were performed. We have enrolled PwPD if the highest level of clinical diagnostic certainty, defined by the term *clinically established Parkinson’s disease* based in accordance with the revised diagnostic criteria of the Movement Disorders Society (MDS) [31]. We assessed demographic data, medical history, the current medication state (to calculate the levodopa-equivalent daily dosage, LEDD) [32,33], and potential exclusion criteria (e.g., the presence of any other neurological comorbidity). Study participants were recruited via our outpatient clinics and from pre-existing cohorts. Trained movement disorders specialists performed standardized clinical assessments (e.g., the Montreal Cognitive Assessment, MoCA) and a neurological examination following the MDS-Unified Parkinson’s Disease Rating Scale subscore III (MDS-UPDRS-III) [34,35]. The assessment of motor symptoms in PwPD was performed after a 12-hour withdrawal of symptomatic treatments.

### Neuroimaging acquisition

We performed multimodal neuroimaging at a 3T *Philips Ingenia* scanner. T_1_-weighted-neuroimaging was performed with a sagittal multi-slice T_1_-weighted Multi-Echo Magnetization Prepared - RApid Gradient Echo (MRMPRAGE) sequence with the following protocol parameters: TR/TE1/ΔTE: 14/2.1/3.1 ms. T_I_ delay: 897.62 ms; flip angle: 9°; 1 × 1 × 1 mm^3^ resolution; 240 × 240 mm^2^ field of view; 180 slices without gap; turbo field echo factor: 120; compressed sensing factor: 6; total scan time: 2 minutes and 54 seconds.

CEST images were acquired using a 0.95 s continuous wave saturation with a nominal B_1_ field strength of 0.8 μT. A 3D Gradient- and Spin-Echo (GRASE) readout was used with TE/TR = 13 ms/2.5 s, FOV = 220 × 220 × 130 mm³, and spatial resolution = 3.5 × 3.5 × 5 mm³. The image matrix was resized to 96 × 96 during reconstruction. 53 offsets within the range of 8 to −2.2 ppm were used in this study, with a scan time of 8 minutes 50 seconds.

T_1_ maps were obtained using the dual flip angle (DFA) method with FOV = 220 × 220 × 140 mm³, and spatial resolution = 2.3 × 2.3 × 5 mm³, excluding the top and bottom slices to match the CEST images, TR/TE = 25/2 ms and flip angles of 5° and 30°. The scan time was 34 seconds per flip angle.

### Neuroimaging preprocessing and analyses

The pipeline is summarized in Figure 1. Both GuanCEST and amideCEST signals were extracted simultaneously using a polynomial and Lorentzian fitting (PLOF) method as previously described [21,36,37]. The background spectrum was fitted using polynomial functions, and the residual Z-spectrum, after background subtraction, was fitted with Lorentzian functions to quantify the CEST signals. In-house Matlab R2024a (Natick, Massachusetts) codes were used for all the evaluation mentioned above.

**Figure 1.**
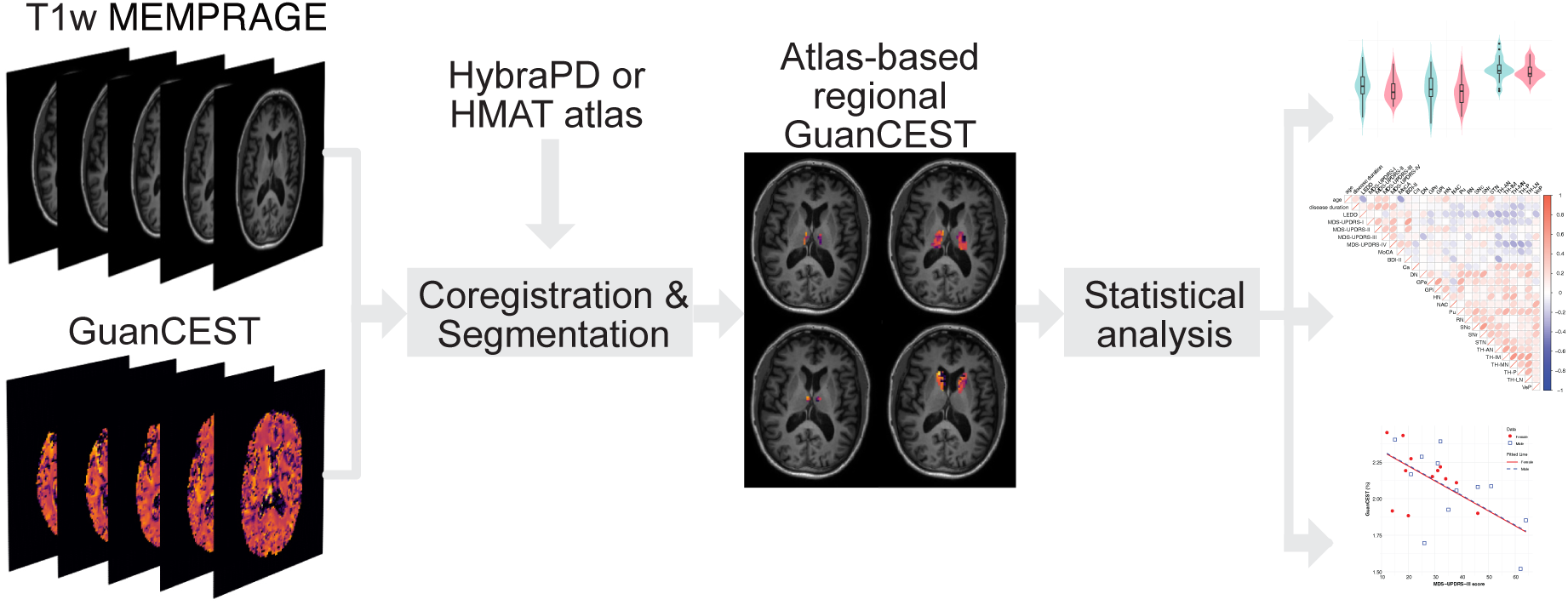
Data acquisition, preprocessing and statistical analyses pipeline. After extracting the GuanCEST maps from the raw CEST data, we segment them with the help of high-resolution T1-weighted (T1w) Multi-Echo Magnetization Prepared - RApid Gradient Echo (MRMPRAGE) images and two different PD-specific atlases (HybraPD and HMAT). Based on the regional GuanCEST levels outliers are removed, followed by statistical analysis (Student’s t-test, Kendall’s correlation, and ANCOVA). GuanCEST, guanidino chemical exchange saturation transfer MRI.

We performed ROI analyses focusing on neuroanatomical regions implicated in PD pathophysiology [38,39], particularly those associated with motor symptoms. T1-weighted MEMPRAGE images were first co-registered to an atlas using FSL (version 6.0.7.1) [40–42], followed by rigid registration to the CEST M0 image [43,44]. The combined transformation matrices were used to align atlas-defined ROIs to the CEST image space. HybraPD [38] atlas is a Parkinson’s disease-specific human brain atlas, which delimitates subcortical structures, with a focus on regions primarily associated with the motor symptoms of PD. These subcortical nuclei included: putamen (Pu), caudate nucleus (Ca), nucleus accumbens (NAC), ventral pallidum (VeP), internal and external globus pallidus (GPi and GPe), pars reticulata and pars compacta of substantia nigra (SNr and SNc), red nucleus (RN), subthalamic nucleus (STN), habenular nuclei (HN), and thalamus (TH). The thalamus was further segmented into 5 sub-regions, that is, anterior nuclei (TH-AN), median nuclei (TH-MN), internal medullary lamina (TH-IML), lateral nuclei (TH-LN), and pulvinar (TH-P). And the HMAT [39] atlas includes six sensorimotor regions: pre-supplementary motor area (preSMA), supplementary motor area proper (SMA), dorsal premotor cortex (PMd), ventral premotor cortex (PMv), primary motor cortex (M1), and primary somatosensory cortex (S1). For the HCs, the average combines both the left and right side; for the PwPD, only the hemisphere responsible for the clinically affected side has been considered. All segmentations have been manually reviewed (by J.P.) prior to further analysis. Co-registration and segmentation were conducted using in-house Python (version 3.9.17) code.

An average difference map of GuanCEST between HC and PwPD was generated by normalizing the subtraction between the two groups, shown in Figure 2. Each subject’s GuanCEST map was upsampled and coregistered to the MNI average brain atlas. The HC average map was computed by averaging all HC subjects, while the PwPD average map was constructed by averaging the left hemisphere of right-side-affected PwPD and the right hemisphere of left-side-affected PwPD. Cerebrospinal fluid (CSF) regions were excluded using an intensity thresholding method based on the MNI atlas. All processing steps were performed using MATLAB and Python codes as previously described.

**Figure 2.**
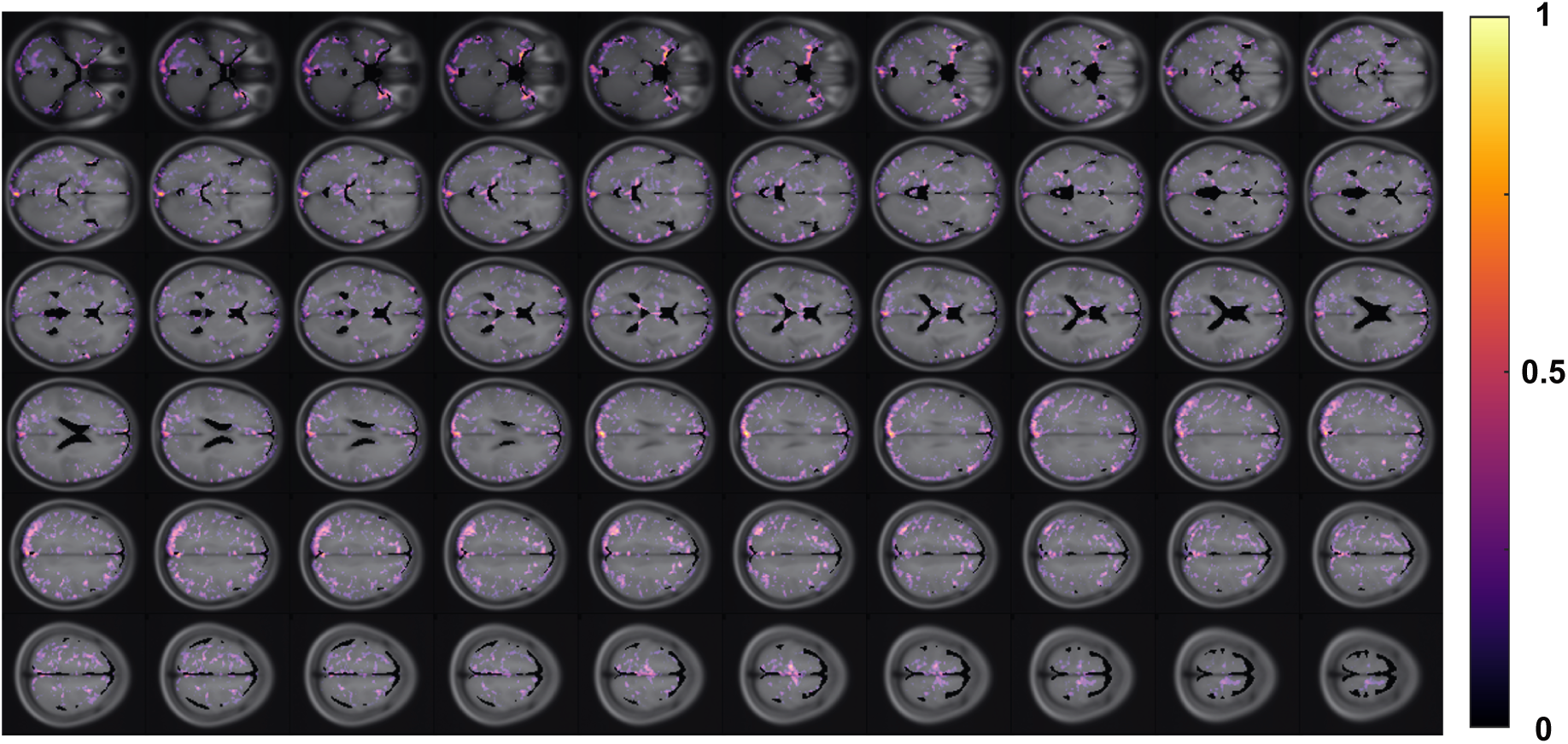
Group-averaged difference in GuanCEST across the brain. The difference (HC – PwPD) in GuanCEST values is averaged across all subjects. For the control group (HC), values represent the average from all HC participants. For the PwPD group, the left cerebrum values are averaged from right-side-affected individuals, and the right cerebrum values are averaged from left-side-affected individuals. The resulting difference map is normalized to a 0–1 scale and overlaid on the MNI T1-weighted structural image atlas, showing the middle 60 slices of interest. Higher values indicate a greater GuanCEST reduction in PwPD relative to HC in that region.

### Statistics

Group differences between PwPD and HCs in each region were evaluated using a two-sided Student’s t-test, with a p-value of less than 0.05 considered significant. The correlations between all covariates—age, gender, disease duration, LEDD, MDS-UPDRS I-IV and total scores, Hoehn and Yahr stage, MoCA, BDI-II, and GuanCEST values in each subregion (as defined by two atlases)—were calculated using Kendall’s correlation for each atlas. Kendall’s correlation is a non-parametric method that does not assume an underlying data distribution and can deal with ties. P-values for the correlations were also computed.

The analysis of covariance (ANCOVA) was performed in the PwPD group to quantify the relationship between GuanCEST values and disease severity, accounting for age and gender. Only significant pairs from the Kendall’s correlation analysis were examined in this step. Disease severity was reflected by using disease duration and MDS-UPDRS-III score as separate outcomes. If significant findings were observed in the GuanCEST values of specific regions (based on Student’s t-test, ANCOVA, or Kendall’s correlation), we similarly examined the amideCEST values and correlations in those regions to determine if significant differences were also present for amideCEST.

Descriptive statistics are reported as mean ± standard deviations (SD) except for Hoehn and Yahr stage, which is reported as median (minimum; maximum). Statistical analyses were done with R (version 4.4.1) on a Macintosh workstation (macOS Sonoma, version 14.6.1). Graphs were created using GraphPad Prism (version 10.3.1).

## RESULTS

### Demographics and clinical data

We enrolled 25 PwPD (12 females, 13 males, age: 67.7 ± 7.1 years) and 24 age- and sex-matched HCs (10 females, 14 males, age: 65.4 ± 7.5 years). The PwPD were in a midstage disease phase with a mean disease duration of 8.2 ± 4.8 years, an overall moderate motor-(MDS-UPDRS-III score: 31.2 ± 14.0 points, MDS-UPDRS-IV score: 2.2 ± 2.3 points, Hoehn and Yahr scale: 2 (1;4)) and non-motor (MDS-UPDRS-I score: 9.4 ± 4.8 points, MDS-UPDRS-II score: 8.6 ± 5.5 points) symptom burden. There were no systematic differences between PwPD and HCs with respect to cognitive impairment (MoCA score: 28.1 ± 1.54 points for PwPD, and 27.8 ± 2.40 points for HCs; p = 0.72). The PwPD had higher depression scores (BDI-II: 7.5 ± 5.0 points for PwPD, 2.7 ± 5.7 for HCs; p = 0.006). All demographic and clinical data are listed in Table 1.

**Table 1.** Demographic and clinical characteristics of the study cohort.

|  | **PwPD (n = 25)** | **HCs (n = 24)** |
| --- | --- | --- |
| **Sex [f/m]** | 12/13 | 10/14 |
| **Age [years]** | 67.7 ± 7.1 | 65.4 ± 7.5 |
| **Disease duration [years]** | 8.2 ± 4.8 | N/A |
| **MDS-UPDRS-I** | 9.36 ± 4.78 | N/A |
| **MDS-UPDRS-II** | 8.56 ± 5.53 | N/A |
| **MDS-UPDRS-III** | 31.24 ± 14.04 | N/A |
| **MDS-UPDRS-IV** | 2.24 ± 2.31 | N/A |
| **MDS-UPDRS (total)** | 51.40 ± 20.55 | N/A |
| **Hoehn and Yahr stage\*** | 2 (1;4) | N/A |
| **LEDD [mg/d]** | 966.0 ± 458.2 | N/A |
| **MoCA** | 28.12 ± 1.54 | 27.79 ± 2.40 |
| **BDI-II** | 7.48 ± 4.97 | 2.67 ± 5.65 |

All metrics are reported as mean ± standard deviation. \*median (minimum; maximum). BDI-II: Beck's Depression Inventory, version 2. f: female. HCs: healthy controls. LEDD: levodopa-equivalent daily dosage. m: male. MDS-UPDRS: Movement Disorders Society - Unified Parkinson's Disease Rating Scale. MoCA: Montreal Cognitive Assessment. N/A: not applicable. PwPD: patients with Parkinson's disease.

### GuanCEST mapping

The group-averaged difference map of GuanCEST between HCs and PwPD is shown in Figure 2. Positive values reflect lower GuanCEST signal in PwPD relative to HCs, with reductions distributed across the brain. More pronounced differences were observed in the deep gray matter and adjacent cortical areas. Representative whole-brain GuanCEST maps from individual PwPD and HC participants are presented in Supplementary Figure 1. Regional GuanCEST values for both groups are plotted in Figure 3A and detailed in Supplementary Table 1. Although a general trend of lower GuanCEST values in PwPD was observed, only the caudate nucleus showed a statistically significant reduction (PwPD: 1.67 ± 0.26% vs. HCs: 1.82 ± 0.16%; p = 0.023). To improve contrast-to-noise ratio, smaller regions in the thalamus (TH-AN, TH-MN, TH-IML, TH-LN, TH-P), basal ganglia (Ca, GPe, GPi, Pu, NAC, VeP), and a composite deep gray matter region (sum of all HybraPD subregions) were grouped for analysis. However, no additional significant group differences were identified (all p-values between 0.6 and 0.8, data not shown). Representative amideCEST maps from the same HC and PwPD participants are shown in Supplementary Figure 2. Group comparisons based on HybraPD regions are presented in Figure 3B, revealing no significant regional differences in amideCEST values. Violin plots comparing HMAT-based GuanCEST and amideCEST values are also shown in Supplementary Figure 3.

**Figure 3.**
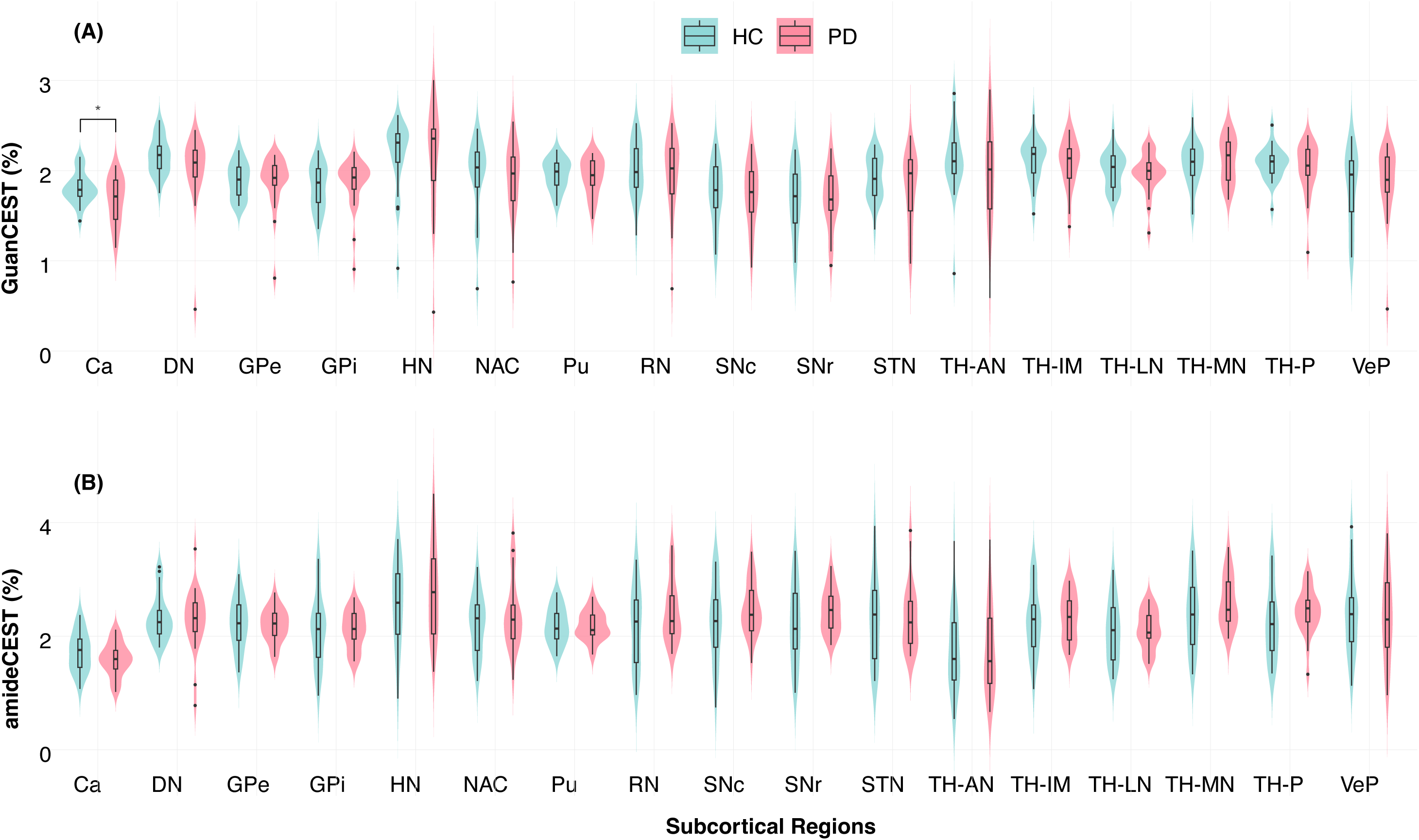
Group comparison of (A) GuanCEST and (B) amideCEST levels in subcortical regions based on the HybraPD atlas. The violin plot shows the distribution of GuanCEST levels across groups, with the box representing the interquartile range (IQR), the middle short bar indicating the median, and the extending black lines from the box defined as the first quartile minus 1.5 times the IQR and the third quartile plus 1.5 times the IQR, respectively. Isolated dots represent outliers, and * denotes p < 0.05 according to Student’s t-test. Ca: caudate nucleus, DN: dentate nuclei, GPe: external globus pallidus, GPi: internal globus pallidus, HN: habenular nuclei, NAC: nucleus accumbens, Pu: putamen, RN: red nucleus, SNc: pars compacta of substantia nigra, SNr: pars reticulata of substantia nigra, STN: subthalamic nucleus, TH-AN: anterior nuclei of thalamus, TH-IML: internal medullary lamina of thalamus, TH-MN: median nuclei of thalamus, TH-P: pulvinar of thalamus, TH-LN: lateral nuclei of thalamus, VeP: ventral pallidum. GuanCEST, guanidino chemical exchange saturation transfer MRI.

### Relationship between GuanCEST signals and clinical characteristics

In the PwPD cohort, we examined Kendall’s correlations between GuanCEST values and clinical variables—including age, disease duration, MDS-UPDRS-III score, and LEDD—in subcortical and sensorimotor cortical regions defined by the HybraPD (Supplementary Figure 4) and HMAT atlases (Supplementary Figure 5), respectively. GuanCEST values in the TH-IML showed a significant negative correlation with MDS-UPDRS-III scores (r = −0.44, p = 0.03), and a non-significant but notable trend with disease duration (r = −0.35, p = 0.09). A similar trend was observed in the TH-LN with MDS-UPDRS-III scores (r = −0.39, p = 0.06). The SNr showed a positive but non-significant correlation with disease duration (r = 0.35, p = 0.10). In contrast, no significant correlations were found in sensorimotor cortical regions defined by HMAT (all p > 0.1), although most showed a negative trend. Notably, GuanCEST signal levels in these cortical regions were consistently lower than even the lowest values observed in subcortical regions, suggesting a potential floor effect that may limit detection of clinical associations.

ANCOVA analysis was conducted solely for regions showing significant correlations between GuanCEST values and clinical data. Figure 4A presents the regression between GuanCEST values in the TH-IML and the MDS-UPDRS-III score for females and males separately, adjusting for age and sex. GuanCEST in the TH-IML decreases significantly by approximately 0.01% for each point increase in the MDS-UPDRS-III score (p = 0.008). On the contrary, amideCEST doesn’t show a significant change with the MDS-UPDRS-III score (p=0.22), when regressed with age and sex (Fig. 4B). Similar findings of GuanCEST in TH-LN are presented in Supplementary Figure 7A, demonstrating a significant decrease as the disease progresses (p=0.014). In contrast, the amideCEST signal in TH-LN doesn’t show a significance change (p=0.27), as shown in Supplementary Figure 7B.

**Figure 4.**
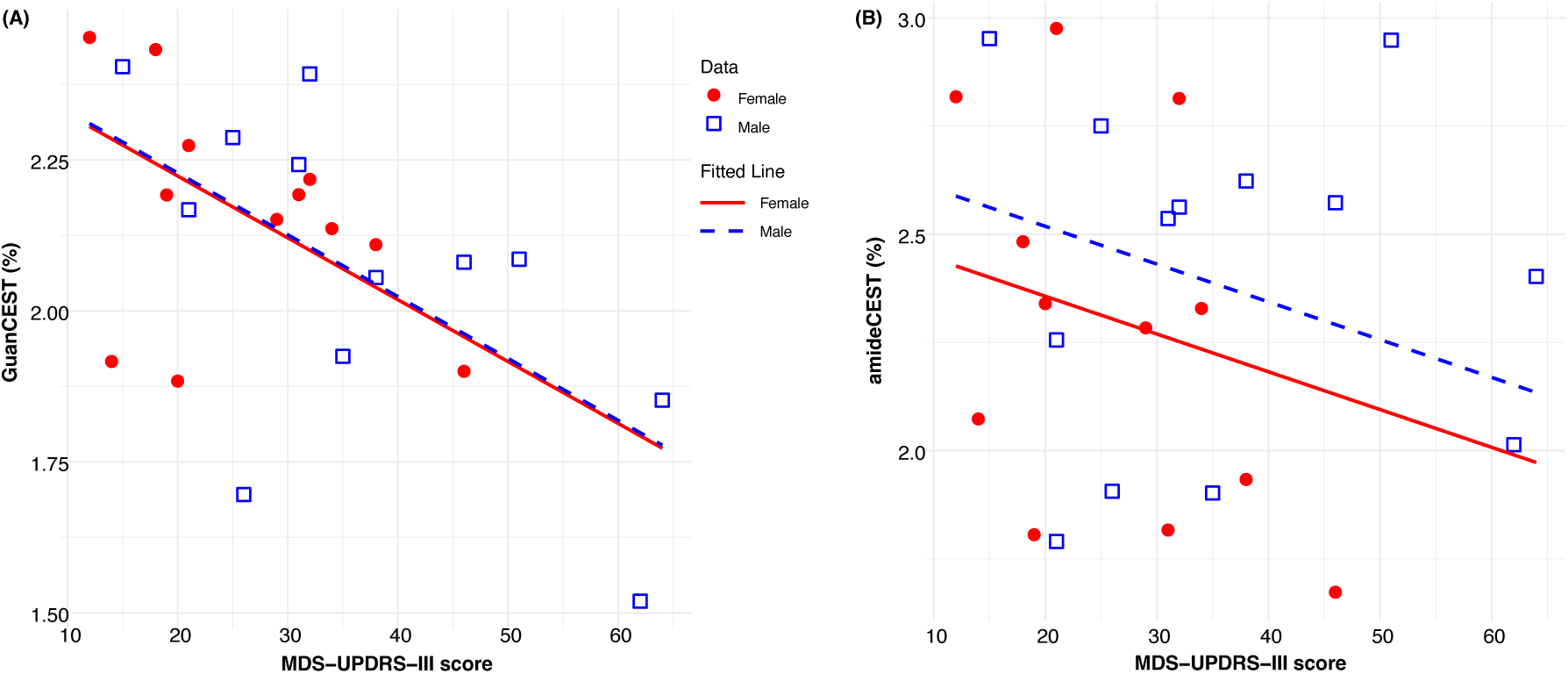
Analysis of covariance results for Guanidino chemical exchange saturation transfer (GuanCEST) signal intensity levels in the internal medullary lamina of the thalamus, compared with amide CEST (amideCEST). (A) The correlation between GuanCEST values and MDS-UPDRS-III score is modeled as a linear function, accounting for age and sex, with separate fits for males (blue squares represent individual data points, blue dashed line for fitted trend) and females (red circles for individual data points, red solid line for fitted trend). The fitted lines assume a fixed age of 67.7 years, which corresponds to the average age of patients with PD. GuanCEST in the internal medullary lamina of the thalamus decreases significantly by approximately 0.01% for each point increase in the MDS-UPDRS-III score (p = 0.008). (B) Similar correlation between amideCEST and MDS-UPDRS-III score accounting for age and sex. The decrease of amideCEST per increase of MDS-UPDRS-III score is not significant (p = 0.22). MDS-UPDRS: Movement Disorders Society - Unified Parkinson’s Disease Rating Scale. GuanCEST, guanidino chemical exchange saturation transfer MRI.

### Evaluation of amideCEST shows no significant changes in protein profile and pH

In regions where significant differences (based on Student’s t-test) or correlations (based on Kendall’s correlation) were found for GuanCEST—specifically in the TH-LN, the TH-IML, and the Ca—amideCEST values were also examined similarly as described above. In the Ca, amideCEST is found to decrease in PwPD, averaging (1.60 ± 0.29%) compared with HCs, who average (1.72 ± 0.35%), although this difference is not statistically significant (p = 0.18). Additionally, no significant Kendall’s correlations were observed between the TH-IML, TH-LN, SNc, and clinical information such as the MDS-UPDRS III scores or the disease duration (Supplementary Figure 6).

## DISCUSSION

In this study, we applied GuanCEST MRI to capture Cr-weighted signals in brain regions previously implicated in PD. While GuanCEST values tended to be lower in PwPD compared to controls across most regions, a significant group difference was observed only in the caudate nucleus. Within the PwPD group, GuanCEST signals in thalamic subregions were significantly associated with disease severity, as measured by both MDS-UPDRS III scores and disease duration. The absence of significant group differences in thalamic regions between PwPD and HCs, despite correlations with clinical severity within PwPD, may reflect heterogeneity in disease progression and highlights the importance of accounting for individual variability. The wide range of GuanCEST values among PwPD underscores the need for within-group analyses in studying bioenergetic markers. Although GuanCEST reflects combined contributions from CrCEST and ArgCEST, ArgCEST arises from mobile cellular proteins. In this study, amideCEST— used as a surrogate for mobile protein content—showed no significant group differences or associations with disease severity, suggesting minimal changes in protein composition. Assuming ArgCEST contributions are negligible, the observed GuanCEST reductions are likely attributable to Cr changes. Notably, other studies have reported altered protein-sensitive imaging signals in PD using APTw MRI [20]. However, APTw includes a mix of saturation transfer effects and lacks specificity [25,45], which may account for the discrepancies. Overall, while our findings support the potential of GuanCEST MRI for detecting Cr-related alterations in PD, further methodological validation is needed to confirm its specificity and clinical utility. The caudate nuclei showed a significant decrease in GuanCEST in PwPD, which, to our knowledge, has not yet been observed with other neuroimaging-based approaches. Reduced Cr levels in the Ca of PwPD may result from the region’s selective vulnerability to PD-specific pathology, including dopaminergic degeneration, mitochondrial dysfunction, and altered energy metabolism. The high metabolic demands of the Ca and its sensitivity to neuroinflammation and α-synuclein accumulation could uniquely disrupt Cr production compared to other brain regions. Additionally, regional differences in glial function, blood-brain barrier integrity, or compensatory mechanisms may further contribute to this localized metabolic change. The meta-analysis of nine 31P MRS studies in PD (n = 183) [16] revealed a decreased phosphomonoester (PME) to phosphodiester (PDE) ratio, corresponding to reduced phosphocholine and phosphoethanolamine relative to glycerophosphocholine and glycerophosphoethanolamine. This finding suggests a potential breakdown of the phospholipid membrane, which might result from the mitochondrial dysfunction. Furthermore, leave-one-out analyses indicated a significant reduction in high-energy metabolite ratios (PCr/Pi and ATP/Pi) in the temporal-parietal lobe. Another 31P MRS study of 30 PwPD and 25 HCs reports a remarkable decrease of high-energy phosphate (PCr/Pi, aATP/Pi, and (aATP+PCr)/Pi) in the patients, particularly in the basal ganglia [46]. These results support the notion that ATP production becomes impaired in the presence of severe mitochondrial dysfunction, leading to reduced PCr regeneration. Since Cr exists in equilibrium with PCr, prolonged PCr depletion may result in Cr depletion as well, a phenomenon that could explain progressive energy failure in advancing PD. Notably, the Ca is affected later than the putamen, which is already severely impacted by the time PD is clinically established. The lower GuanCEST levels in Ca may reflect surviving neurons depleting creatine as an energy buffer to compensate for metabolic demands. This may even suggest that Ca represents the “leading edge” of neurodegeneration in the striatum in this cohort.

The GuanCEST signal in the TH-IML and TH-LN decreases in correlation with disease severity. Interestingly, the TH-IML is integral to motor networks, serving as a key structural pathway that facilitates communication between motor-related thalamic nuclei, including the TH-LN, and their connections to the basal ganglia and motor cortex [47]. This network coordination is crucial for motor control, but the TH-IML’s dense, metabolically active fibre architecture may render it particularly susceptible to bioenergetic deficits. Impaired Cr metabolism in the TH-IML could disrupt thalamocortical and thalamobasal ganglia pathways, contributing to the motor symptom severity of PwPD. Furthermore, the TH-LN, which are also connected through the TH-IML and would involve the ventral posterolateral nucleus, the major relay for motor control through the basal ganglia-thalamo-cortical network, may experience metabolic impairments that underly motor dysfunction. GuanCEST MRI could help detect these bioenergetic vulnerabilities, providing insights into the role of the TH-IML and TH-LN in PD progression and justify potential targets for intervention.

Given the critical role of Cr in the central nervous system, numerous preclinical and clinical studies have examined its potential in treating neurodegenerative disorders, including PD [48,49]. For instance, in PD mouse models with mitochondrial dysfunction induced by peritoneal injection of the neurotoxin and complex I inhibitor 1-methyl-4-phenyl-1,2,3,6-tetrahydropyridine (MPTP), pre-treatment with a diet containing 1% Cr for two weeks provided nearly complete protection of dopaminergic neurons compared to a control diet [50]. Furthermore, in a chronic subcutaneous MPTP mouse model, combining Cr with coenzyme Q_10_ offered increased neuroprotection, demonstrated by reduced lipid peroxidation damage and decreased alpha-synuclein accumulation in the STN’s dopaminergic neurons [51]. Clinical trials, however, have shown inconsistent results, with two meta-analyses on this topic [52,53] largely refuting the therapeutic effect of Cr, as evidenced by two large-cohort double-blinded randomized clinical trials [54,55]. This variability may be due, in part, to suboptimal Cr dosing strategies, the lack of target engagement methods, and the absence of pathophysiology-based patient stratification. Given the broad heterogeneity in suspected mechanisms of both disease initiation and progression indicated by a variety of genetic and environmental factors, including neuroinflammation, genetic predisposition to energetic deficits, α-synuclein templating, and lysosomal function, it is not surprising that a one-size-fits all approach for disease modification has failed in clinical trials for PD. The contrast between preclinical and clinical findings underscores the complexity of Cr’s role in PD and suggests a need for personalized and precision-based approaches in treatment based on predominant pathophysiologic mechanisms at the individual level.

This study has several limitations:

(1) *Sample size*: The relatively small cohort limited statistical power to detect subtle effects. Given the substantial interindividual variability observed among PwPD, larger studies are needed to better characterize Cr-related changes.
(2) *Specificity of GuanCEST*: While amideCEST was used as a surrogate to estimate Arg contributions within the GuanCEST signal, the influence of other mobile proteins and potential pH variations cannot be excluded. More advanced imaging approaches are needed to improve specificity for Cr.
(3) *Spatial resolution*: The resolution of GuanCEST imaging was adequate for larger structures but limited in smaller subcortical regions, where partial volume effects may reduce signal specificity. Improved resolution or post-processing methods may enhance localization but may require trade-offs in scan time or signal quality.
(4) Multiple comparisons: ROI-based analyses were not corrected for multiple comparisons, raising the possibility of type I error. However, regions were pre-selected based on established relevance to PD pathology, supporting the exploratory value of these findings and guiding future research.

## CONCLUSIONS

In conclusion, we utilized Cr-weighted GuanCEST signals to investigate regional changes in PwPD compared with age- and sex-matched HCs. While a decrease in GuanCEST signal was observed across most gray matter regions, only the Ca showed a statistically significant reduction. Further correlation analysis revealed significant associations between GuanCEST Cr-weighted signals in thalamic regions and the individual disease severity metrics, as indicated by the disease duration and the MDS-UPDRS III scores.

## Data Availability

All data produced in the present study are available upon reasonable request to the authors

## ACKNOWLEDGMENT

We kindly thank our patients and their caregivers for participating in this study. The National Institutes of Health supported this project through grants P41EB031771.

## AUTHOR ROLES

KW: design, analysis, writing, and editing of the final version of the manuscript.

NNY: design, execution, analysis, writing, and editing of the final version of the manuscript.

ZY: analysis, writing, and editing of the final version of the manuscript.

TMD: design, and editing of the final version of the manuscript

PvZ: design, execution, analysis, writing, and editing of the final version of the manuscript.

KAM: design, execution, analysis, writing, and editing of the final version of the manuscript.

JX: design, analysis, writing, and editing of the final version of the manuscript.

JP: design, execution, analysis, writing, and editing of the final version of the manuscript.

## Notes

### Competing Interest Statement

The authors have declared no competing interest.

### Funding Statement

This study was funded by NIH through the grant P41EB031771.

### Author Declarations

The present study has been performed after receiving prior approval from the institutional review board of Johns Hopkins University, Baltimore, MD, USA, (#00371335) following the revised version of the Declaration of Helsinki.

### Summary of Updates

Correct the corresponding author name, and all the affiliations

