## Supplementary for "Creatine-weighted imaging in patients with Parkinson’s disease"

### Supplementary Materials

**Supplementary Table 1. Guanidino chemical exchange saturation transfer (GuanCEST) values# in subcortical and sensorimotor regions as defined by the HybraPD and HMAT atlas.**

| **Atlas** | **Region** | **PwPD** (n = 25) | **HCs** (n = 24) | **p-value** |
|---|---|---|---|---|
| **HybraPD** | Ca | 1.67 ± 0.26 | 1.82 ± 0.16 | 0.023* |
| | DN | 2.09 ± 0.21 | 2.16 ± 0.20 | 0.244 |
| | GPe | 1.95 ± 0.16 | 1.89 ± 0.19 | 0.273 |
| | GPi | 1.92 ± 0.16 | 1.83 ± 0.25 | 0.104 |
| | HN | 2.23 ± 0.46 | 2.28 ± 0.22 | 0.599 |
| | NAC | 1.95 ± 0.35 | 1.98 ± 0.34 | 0.698 |
| | Pu | 1.94 ± 0.20 | 1.96 ± 0.17 | 0.634 |
| | RN | 2.02 ± 0.33 | 2.02 ± 0.31 | 0.955 |
| | SNc | 1.74 ± 0.35 | 1.75 ± 0.34 | 0.886 |
| | SNr | 1.72 ± 0.30 | 1.67 ± 0.38 | 0.647 |
| | STN | 1.80 ± 0.40 | 1.92 ± 0.25 | 0.213 |
| | TH-AN | 1.96 ± 0.55 | 2.14 ± 0.25 | 0.156 |
| | TH-IML | 2.11 ± 0.23 | 2.14 ± 0.21 | 0.558 |
| | TH-MN | 2.12 ± 0.25 | 2.07 ± 0.25 | 0.502 |
| | TH-P | 2.07 ± 0.21 | 2.08 ± 0.13 | 0.822 |
| | TH-LN | 2.02 ± 0.16 | 2.01 ± 0.20 | 0.824 |
| | VeP | 1.95 ± 0.26 | 1.81 ± 0.42 | 0.188 |
| **HMAT** | M1 | 1.18 ± 0.19 | 1.23 ± 0.26 | 0.429 |
| | PMd | 1.12 ± 0.22 | 1.18 ± 0.26 | 0.397 |
| | PMv | 1.48 ± 0.14 | 1.52 ± 0.12 | 0.303 |
| | S1 | 1.24 ± 0.21 | 1.31 ± 0.25 | 0.313 |
| | SMA | 1.56 ± 0.25 | 1.59 ± 0.27 | 0.705 |

| | | | |
|---|---|---|---|
| preSMA | 1.44 ± 0.24 | 1.54 ± 0.32 | 0.231 |

### The GuanCEST signal intensity is reported in %. * indicates a significant group difference determined by a two-sided Student's t-test with $p < 0.05$. All metrics are reported as mean ± standard deviation. Due to the exploratory nature of this study, no statistical correction for multiple testing has been performed. Ca: caudate nucleus, DN: dentate nuclei, GPe: external globus pallidus, GPi: internal globus pallidus, HN: habenular nuclei, NAC: nucleus accumbens, Pu: putamen, RN: red nucleus, SNc: pars compacta of substantia nigra, SNr: pars reticulata of substantia nigra, STN: subthalamic nucleus, TH-AN: anterior nuclei of thalamus, TH-IML: internal medullary lamina of thalamus, TH-MN: median nuclei of thalamus, TH-P: pulvinar of thalamus, TH-LN: lateral nuclei of thalamus, VeP: ventral pallidum. M1: primary motor cortex, PMd: dorsal premotor cortex, PMv: ventral premotor cortex, S1: primary somatosensory cortex, SMA: supplementary motor area, preSMA: pre-supplementary motor area. GuanCEST, guanidino chemical exchange saturation transfer MRI.

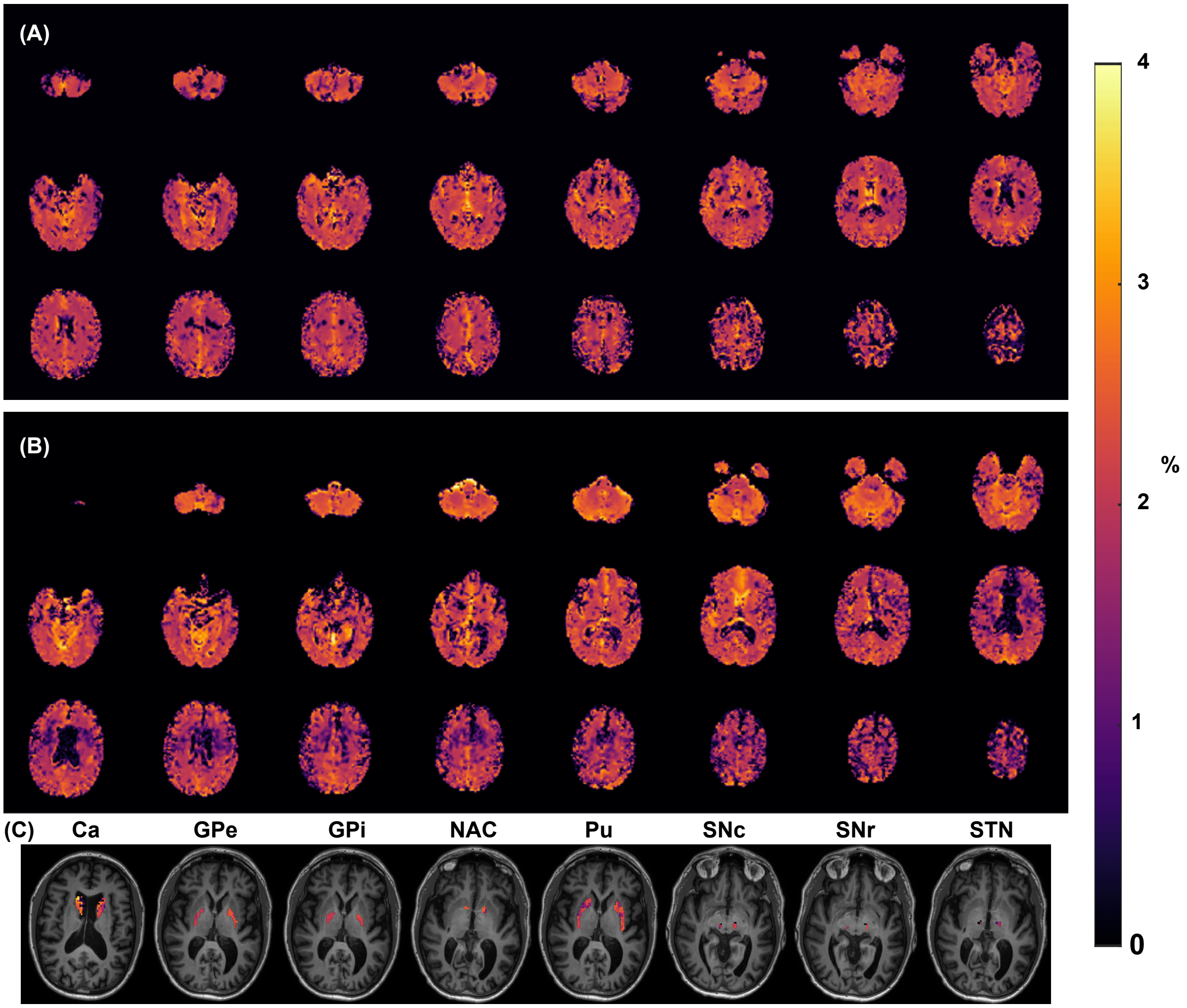


**Supplementary Figure 1. Typical whole-brain GuanCEST maps and segmentation**. Whole-brain GuanCEST maps (middle 24 slices) are shown for a typical HC (A) and a PwPD (B). (C) GuanCEST maps overlaid on T1-weighted MEMPRAGE structural images for selected subcortical regions in a PwPD subject. Ca: caudate nucleus, GPe: external globus pallidus, GPi: internal globus pallidus, NAC: nucleus accumbens, Pu: putamen, SNc: pars compacta of substantia nigra, SNr: pars reticulata of substantia nigra, STN: subthalamic nucleus. GuanCEST, guanidino chemical exchange saturation transfer MRI.

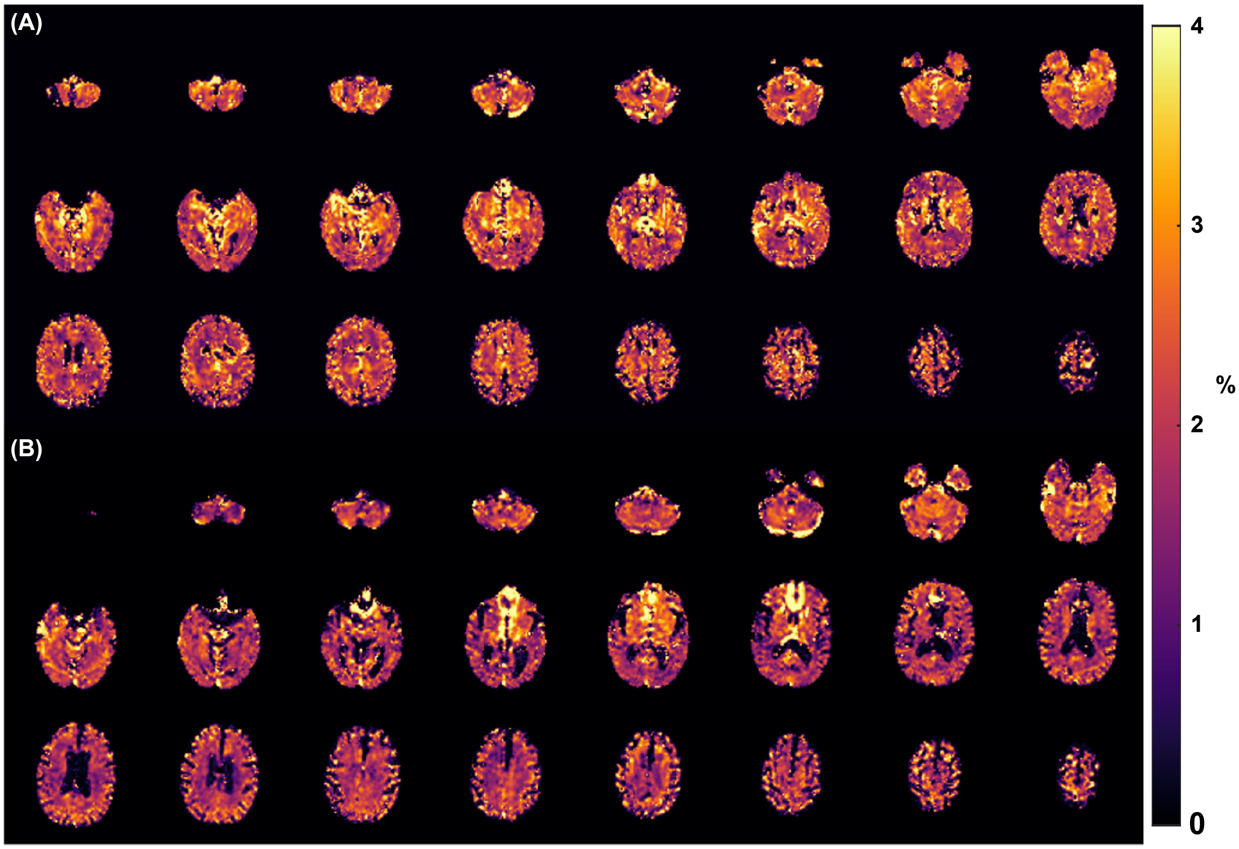


**Supplementary Figure 2. Typical whole-brain amideCEST maps**. Whole-brain amideCEST maps (middle 24 slices) are shown for a typical HC (A) and a PwPD (B).

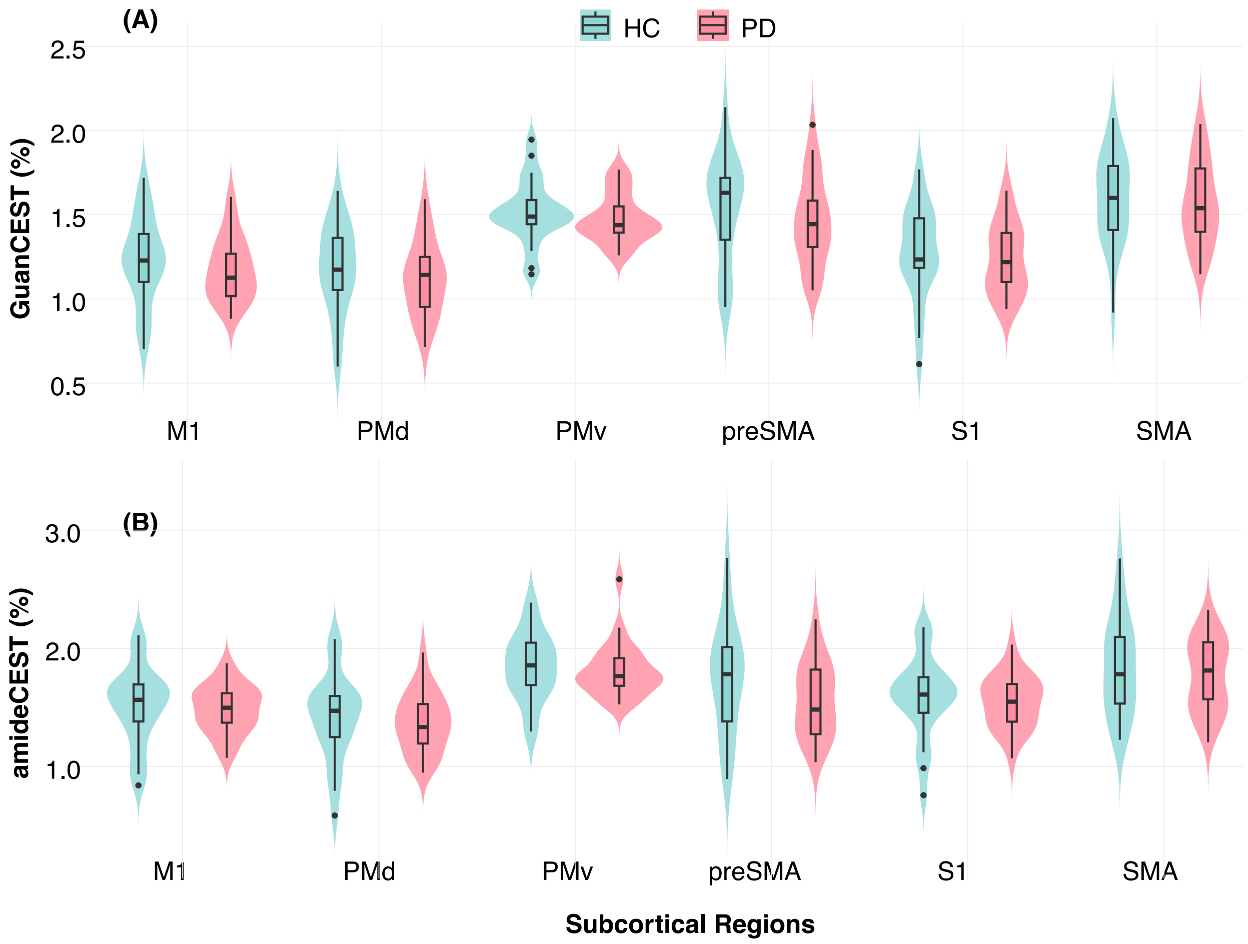


**Supplementary Figure 3. Group comparison of (A) GuanCEST and (B) amideCEST levels in subcortical regions based on the HMAT atlas.** The violin plot shows the distribution of GuanCEST levels across groups, with the box representing the interquartile range (IQR), the middle short bar indicating the median, and the extending black lines from the box defined as the first quartile minus 1.5 times the IQR and the third quartile plus 1.5 times the IQR, respectively. Isolated dots represent outliers. M1: primary motor cortex, PMd: dorsal premotor cortex, PMv: ventral premotor cortex, S1: primary somatosensory cortex, SMA: supplementary motor area, preSMA: pre-supplementary motor area. GuanCEST, guanidino chemical exchange saturation transfer MRI.

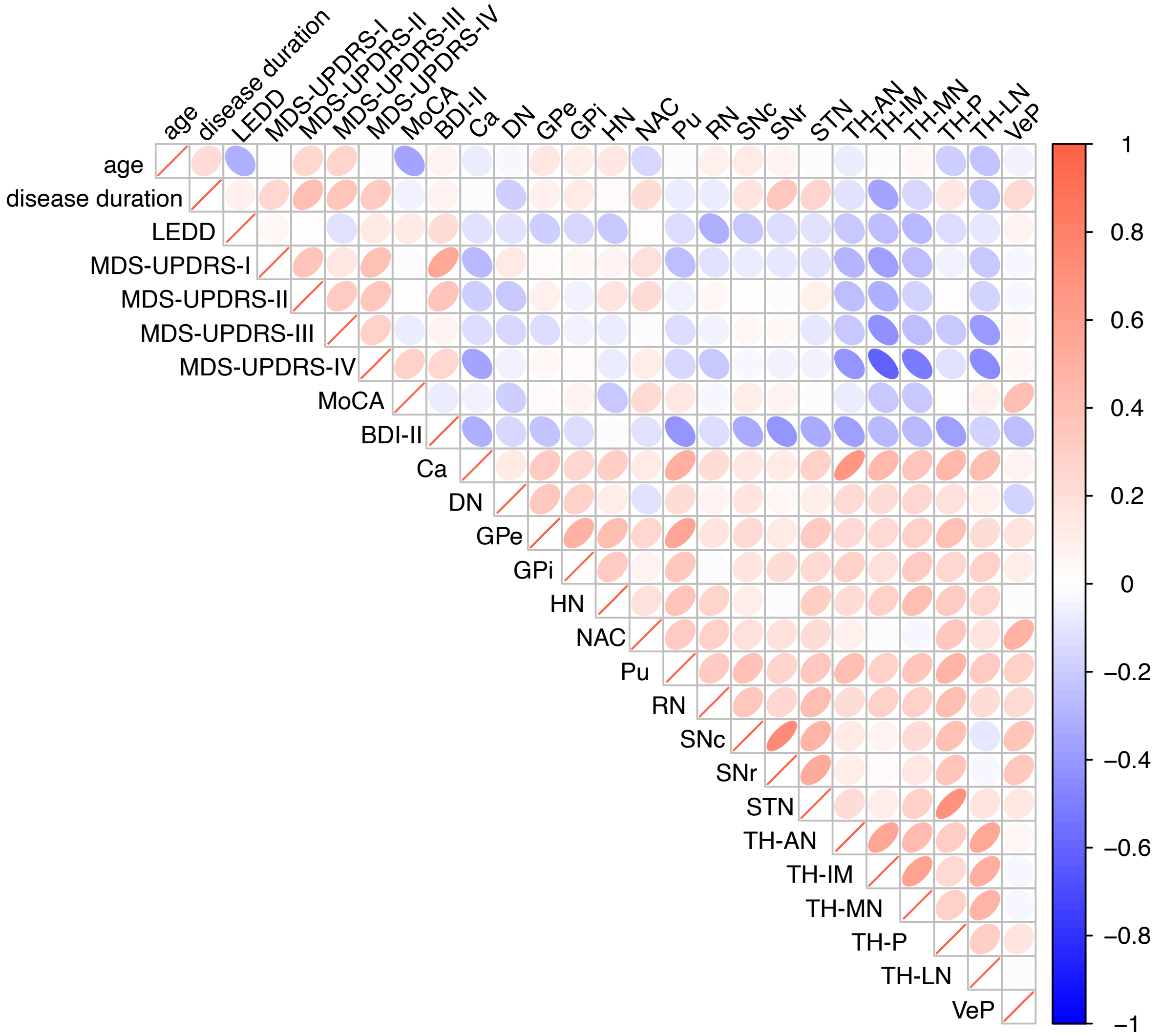


**Supplementary Figure 4. Kendall's correlation between GuanCEST values in subcortical regions (as defined by the HybraPD atlas) and demographic/clinical characteristics.** Ellipses are scaled according to the correlation value, with eccentricity reflecting the strength of the correlation. The color of the ellipses is shaded blue or red, indicating the direction of the correlation, with the intensity of the color representing the magnitude of the correlation. LEDD: levodopa-equivalent daily dosage. MDS-UPDRS: Movement Disorders Society - Unified Parkinson's Disease Rating Scale. MoCA: Montreal Cognitive Assessment. BDI-II: Beck's Depression Inventory, version 2. Ca: caudate nucleus, DN: dentate nuclei, GPe: external globus pallidus, GPi: internal globus pallidus, HN: habenular nuclei, NAC: nucleus accumbens, Pu: putamen, RN: red nucleus, SNc: pars compacta of substantia nigra, SNr: pars reticulata of substantia nigra, STN: subthalamic nucleus, TH-AN: anterior nuclei of thalamus, TH-IML: internal medullary lamina of thalamus, TH-MN:

median nuclei of thalamus, TH-P: pulvinar of thalamus, TH-LN: lateral nuclei of thalamus, VeP: ventral pallidum. GuanCEST, guanidino chemical exchange saturation transfer MRI.

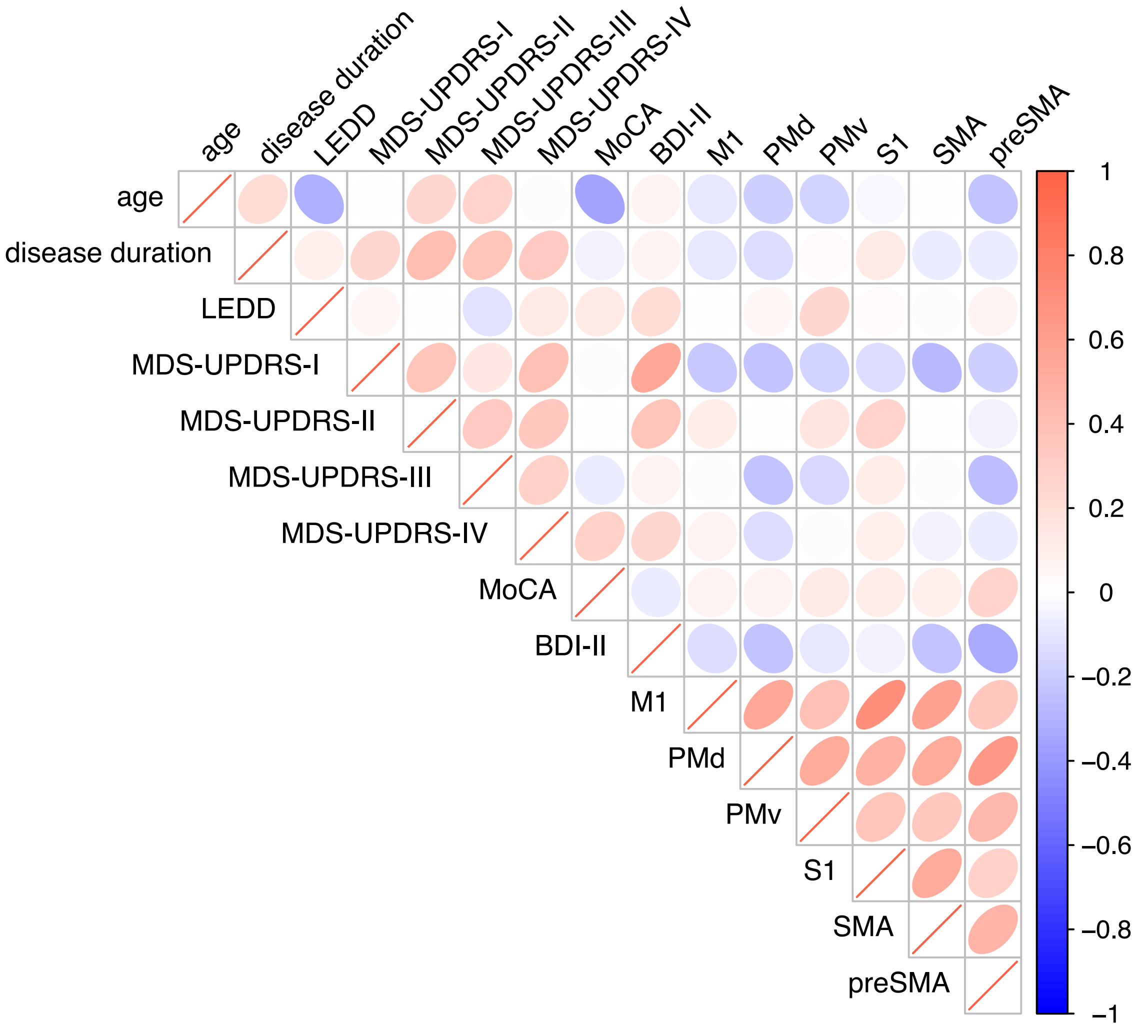


**Supplementary Figure 5. Kendall's correlation between GuanCEST values in subcortical regions defined by HMAT atlas and clinical information.** Ellipses are scaled according to the correlation value, with eccentricity reflecting the strength of the correlation. The color of the ellipses is shaded blue or red, indicating the direction of the correlation, with the intensity of the color representing the magnitude of the correlation. LEDD: levodopa-equivalent daily dosage. MDS-UPDRS: Movement Disorders Society - Unified Parkinson's Disease Rating Scale. MoCA: Montreal Cognitive Assessment. BDI-II: Beck's Depression Inventory, version 2. M1: primary motor cortex, PMd: dorsal premotor cortex, PMv: ventral premotor cortex, S1: primary somatosensory cortex,

SMA: supplementary motor area, preSMA: pre-supplementary motor area. GuanCEST, guanidino chemical exchange saturation transfer MRI.

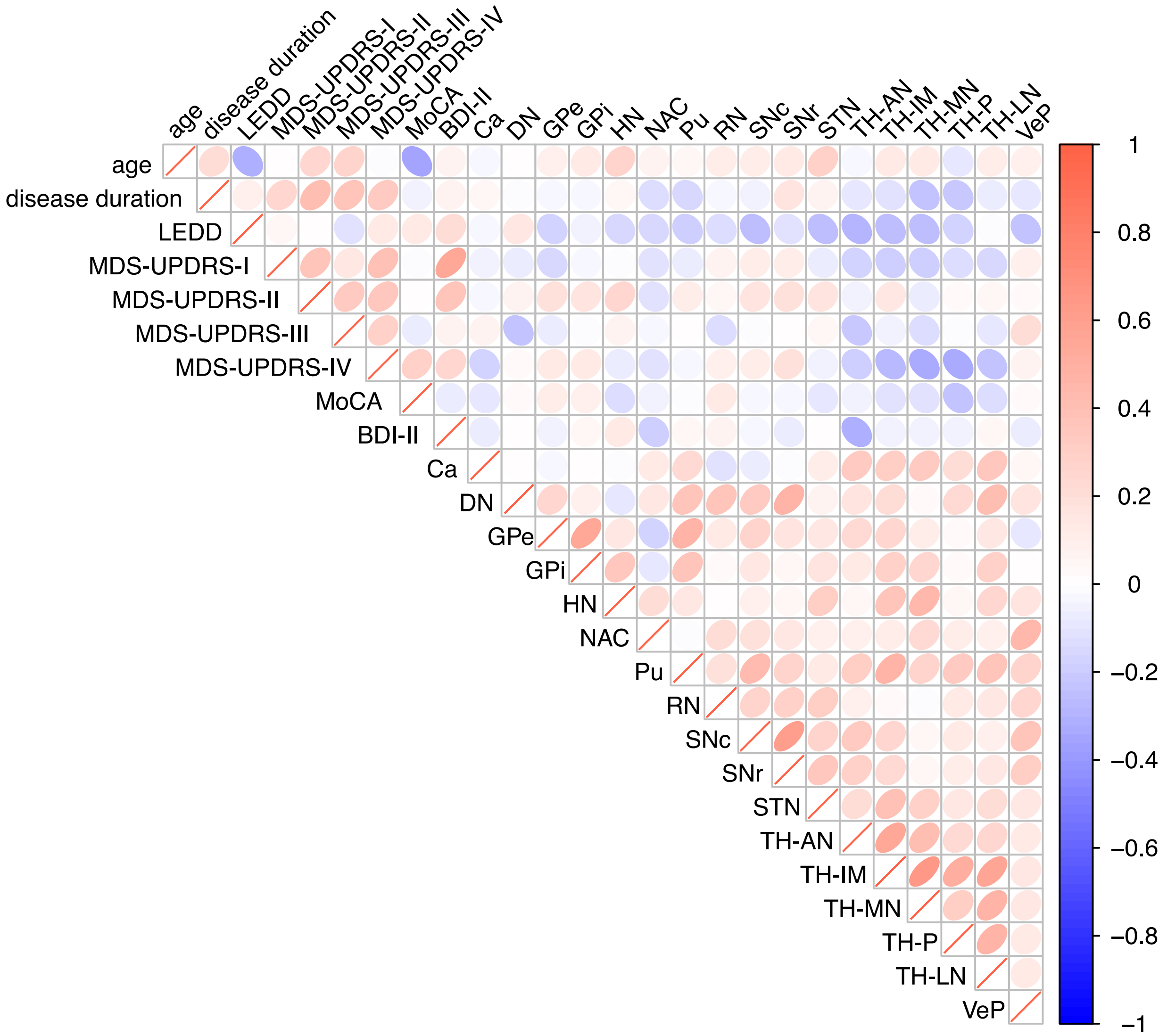


**Supplementary Figure 6. Kendall's correlation between amideCEST values in subcortical regions (as defined by the HybraPD atlas) and demographic/clinical characteristics.** Ellipses are scaled according to the correlation value, with eccentricity reflecting the strength of the correlation. The color of the ellipses is shaded blue or red, indicating the direction of the correlation, with the intensity of the color representing the magnitude of the correlation. Compared with Figure 4, there are less dark blue ellipses between amideCEST values and clinical characteristics. LEDD:

levodopa-equivalent daily dosage. MDS-UPDRS: Movement Disorders Society - Unified Parkinson's Disease Rating Scale. MoCA: Montreal Cognitive Assessment. BDI-II: Beck's Depression Inventory, version 2. Ca: caudate nucleus, DN: dentate nuclei, GPe: external globus pallidus, GPi: internal globus pallidus, HN: habenular nuclei, NAC: nucleus accumbens, Pu: putamen, RN: red nucleus, SNc: pars compacta of substantia nigra, SNr: pars reticulata of substantia nigra, STN: subthalamic nucleus, TH-AN: anterior nuclei of thalamus, TH-IML: internal medullary lamina of thalamus, TH-MN: median nuclei of thalamus, TH-P: pulvinar of thalamus, TH-LN: lateral nuclei of thalamus, VeP: ventral pallidum. amideCEST, amide chemical exchange saturation transfer MRI.

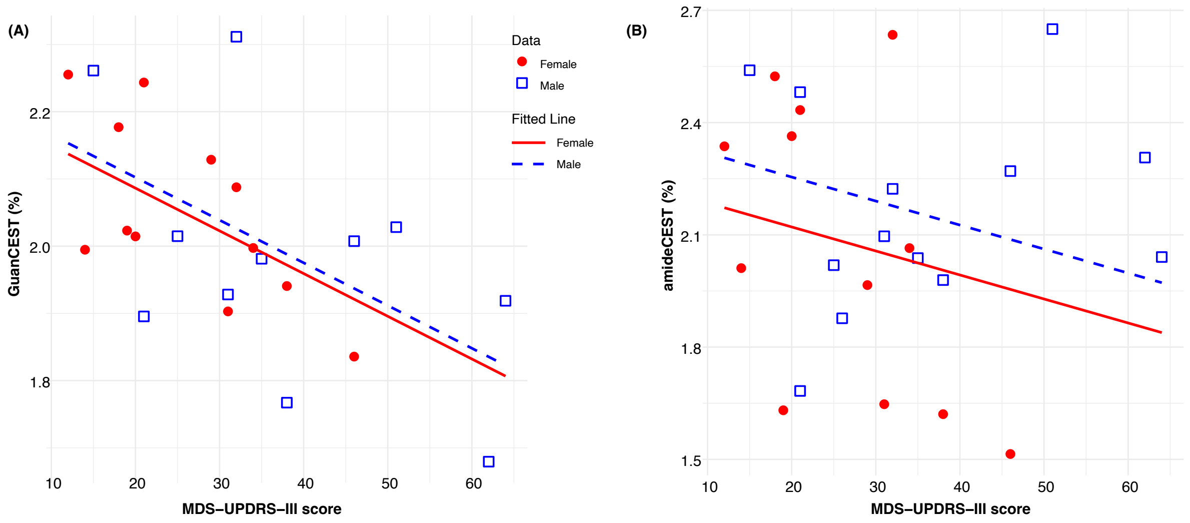


**Supplementary Figure 7. Analysis of covariance results for Guanidino chemical exchange saturation transfer (GuanCEST) levels in the lateral nuclei of the thalamus, compared with amide CEST (amideCEST).** (A) The correlation between GuanCEST values and MDS-UPDRS-III score is modeled as a linear function, accounting for age and sex, with separate fits for males (blue squares represent individual data points, blue dashed line for fitted trend) and females (red circles for individual data points, red solid line for fitted trend). On average, GuanCEST levels in the lateral nuclei of the thalamus decrease by 0.006% per point increase for the MDS-UPDRS-III score (p =

0.014). (B) Similar correlation between amideCEST and MDS-UPDRS-III score accounting for age and sex. The decrease of amideCEST per increase of MDS-UPDRS-III score is not significant ($p = 0.27$). MDS-UPDRS: Movement Disorders Society - Unified Parkinson's Disease Rating Scale. GuanCEST, guanidino chemical exchange saturation transfer MRI.
